# α4β2^*^ Nicotinic Cholinergic Receptor Target Engagement in Parkinson Disease Gait-Balance Disorders

**DOI:** 10.1101/2020.10.11.20210914

**Authors:** Roger L. Albin, Martijn L.T.M. Müller, Nicolaas I. Bohnen, Cathie Spino, Martin Sarter, Robert A. Koeppe, Ashley Szpara, Kamin Kim, Cindy Lustig, William T. Dauer

## Abstract

**Objective:** Attentional function deficits secondary to degeneration of brain cholinergic systems are significant contributors to gait-balance deficits in Parkinson disease (PD). As an initial step towards assessing if α4β2^*^ nicotinic acetylcholine receptor (nAChR) stimulation improves attention and gait-balance function, we assessed target engagement of the α4β2^*^ nAChR partial agonist varenicline.

**Methods:** Non-demented PD participants with cholinergic deficits were identified with [^18^F]fluoroethoxybenzamicol positron emission tomography (PET). α4β2^*^ nAChR occupancy after subacute oral varenicline treatment was measured with [^18^F]flubatine PET. With a dose selected from the receptor occupancy experiment, varenicline effects on gait, balance, and cognition were assessed in a double-masked placebo-controlled crossover study. Primary endpoints were normal pace gait speed and a measure of postural stability.

**Results:** All varenicline doses (0.25 mg per day, 0.25 mg b.i.d., 0.5 mg b.i.d., and 1.0 mg b.i.d.) produced 60% - 70% receptor occupancy. We selected 0.5 mg po b.i.d for the crossover study. Thirty-three (of thirty-four) participants, completed the crossover study with excellent tolerability. Varenicline had no significant impact on the postural stability measure and caused slower normal pace gait speed. Varenicline narrowed the difference in normal pace gait speed between dual task and no dual task gait conditions, reduced dual task cost, and improved performance on a sustained attention test. We obtained identical conclusions in 28 participants in whom treatment compliance was confirmed by plasma varenicline measurements.

**Interpretation:** Varenicline occupied a significant fraction of α4β2^*^ nicotinic acetylcholine receptors, was tolerated well, enhanced attentional function, and altered gait performance. These results are consistent with relevant target engagement. Varenicline or similar agents may be worth further evaluation for mitigation of gait and balance disorders in PD.

## Introduction

Dopamine replacement therapy (DRT)-refractory gait and balance disorders are among the most morbid aspects of Parkinson disease (PD). Gait deficits, including postural instability and freezing, worsen with disease progression and substantially increase fall risk. Falls are a significant source of morbidity in PD patients, with a relatively high rate of serious falls leading to fractures and hospitalizations, precipitation of nursing home placement, and increased mortality associated with falls.^1,2,3,4^

The DRT-refractory nature of gait and postural deficits in PD indicates involvement of non-dopaminergic systems. Considerable evidence suggests that DRT-resistant gait and balance disorders are associated with degeneration of central nervous system (CNS) cholinergic projection systems.^5-13^ Fall risk in PD is likely increased by the conjunction of striatal dopaminergic denervation and degeneration of cholinergic neurons of the basal forebrain corticopetal complex (BFCC) and pedunculopontine-laterodorsal tegmental complex (PPN-LDT). The best defined role of the BFCC is in attention with suggestions that PPN-LDT cholinergic neurons play a role in alertness.^12,14,15^ Preclinical experiments indicate that as BFCC neurons are lost, gait-balance dysfunction may increase markedly as BFCC cholinergic deficits unmask the full impacts of striatal dopaminergic deficits.^12,16^ This model is consistent with results of dual task paradigm experiments, in PD and control participants, indicating that impaired attention is associated with worsening gait-balance functions and increased fall risk.^17^

Cholinergic neurotransmission is mediated by both G-protein coupled receptors and ionotropic nicotinic receptors (nAChRs). The predominant CNS nicotinic receptor is the α4β2^*^ nAChR (^*^potential other subunits).^18^ Stimulation of cortical α4β2^*^ receptors plays an important role in attention and this is likely the mechanism by which nicotine enhances attention. In the setting of BFCC projection degeneration, pharmacologic stimulation of α4β2^*^ nAChRs might improve attention and mitigate gait-balance deficits.

Varenicline (VCN) is a potent (K_i_ = 0.4 nM) α4β2^*^ nAChR partial agonist (efficacy = 45%) used widely for tobacco abuse cessation.^19,20^ VCN has an excellent safety record and favorable pharmacokinetic features.^20-24^ To initiate exploration of the potential of VCN to improve DRT-resistant gait-balance deficits, we performed a target engagement study of VCN in PD participants with neocortical cholinergic deficits. We assessed target engagement along 2 dimensions; VCN binding to brain α4β2^*^ nAChRs, and VCN effects on laboratory-based measures of gait, balance, and cognitive functions.

While there is abundant literature characterizing gait, balance, and fall risk in PD and in normal aging, there are no laboratory-based measures predicting intervention outcomes. In the absence of measures with predictive validity, measures linked to pathophysiologic mechanisms are more likely to be adequate indices of target engagement. In secondary-exploratory analyses, we studied both objective measures of postural sway and gait speed using body-worn inertial sensors and cognitive outcome measures, including a cognitive measure specifically related to disrupted attentional and cholinergic functions.

## Materials and Methods

### Regulatory Compliance

Informed consent was obtained from all participants according to the Declaration of Helsinki. This study was approved by the University of Michigan Medical School Institutional Review Board. An Investigational New Drug application waiver for varenicline study was obtained from the Food & Drug Administration. This study was registered at ClinicalTrials.gov (NCT04403399, NCT02933372).

### Participant Selection

PD participants were recruited from a larger cohort characterized with [^18^F]fluoroethoxybenzovesamicol positron emission tomography ([^18^F]FEOBV PET).^9^ The vesicular acetylcholine transporter ligand [^18^F]FEOBV was used to determine the magnitudes of cortical cholinergic terminal deficits. All participants met the International Parkinson and Movement Disorder Society clinical diagnostic criteria for PD. All underwent [^11^C]dihydrotetrabenazine PET to confirm the presence of characteristic putaminal nigrostriatal dopaminergic terminal deficits. No enrolled participant was demented or was using drugs or supplements with cholinergic properties, or using tobacco products. Because of anecdotal reports of worsening mood disorders, adverse ethanol interactions, and myocardial infarctions, we excluded individuals with an active mood disorder (Geriatic Depression Scale >5 and evidence of recent, worsening mood), alcohol use disorder (Alcohol Use Disorder Identification Test score >7 for those over 65 years; >8 for those 65 years and younger), and active cardiovascular disease. Participants were counseled to avoid alcoholic beverages during study participation. Only participants with cortical cholinergic deficits were enrolled. In PD, the occipital cortex has highest vulnerability for cholinergic transporter losses compared to other brain regions.^25^ Hypocholinergic status was defined as falling within the lower tertile of occipital cortical [^18^F]FEOBV binding in normal older adults. Participants were maintained on stable DRT regimens throughout these experiments. To ensure that there was not a marked difference between VCN interaction with α4β2^*^ nAChRs in PD participant and control brains, we performed a more limited dose-response experiment in normal participants. Age-matched control participants without clinical evidence of parkinsonism or other neurologic disorders, and not using any cholinergic agents or tobacco products, were studied.

### VCN Occupancy of α4β2^*^ nAChRs Study

VCN occupancy of α4β2^*^ nAChRs was assessed with ascending doses of VCN and the selective α4β2^*^ nAChR PET ligand [^18^F]Flubatine.^26^ PD and control participants in the dose response – receptor occupancy study were treated with oral VCN for 10 days with ascending dose schedules. Higher dosing cohorts were begun after the previous dosing cohort completed its scheduled treatment and follow-up. Doses chosen were 0.25 mg per day, 0.25 mg b.i.d., 0.5 mg b.i.d., and 1.0 mg b.i.d. The clinically used VCN dose is 1.0 mg b.i.d. Participants received an initial 0.25 mg dose following confirmation of eligibility and baseline evaluations and were monitored for 4 hours. In participants scheduled for higher VCN doses, the total daily dose was escalated over the next 2 days, followed by 8 days of stable daily VCN dose. α4β2^*^ nAChR agonists may induce nAChR expression, and it is possible that receptor density may not be stable during α4β2^*^ nAChR agonist exposure.^27^ As we cannot measure absolute receptor density, but only relative receptor occupancy, the conventional strategy of imaging participants before and at the end of a drug exposure period might result in underestimation of receptor occupancy. To address this issue, we imaged participants at the end of their drug exposure periods and again after 5 days (∼5 half-lives) of washout from drug exposure. [^18^F]Flubatine was synthesized as described previously.^28^ Participants were scanned on a Biograph TruePoint Model 1094, using a dynamic acquisition of 18 frames over 90 minutes (four x 0.5 min; three x 1 min; two x 2.5 min; two x 5 min; seven x 10 min). α4β2^*^ nAChR occupancy was estimated by comparing [^18^F]Flubatine standardized uptake values (SUVs) on and off VCN. SUVs were calculated as (B-D)/(B-ND), where B is the SUV of the “baseline” scan (off VCN), and D is the SUV of the “drug” scan (on VCN), and ND is estimated from the non-displaceable SUV. ND is calculated from the x-intercept of a regression of (B-D) on D.

### Crossover Study

Following selection of a study dose from the α4β2^*^ nAChR occupancy experiment in PD participants (**Results** below), we completed a double-masked, placebo-controlled crossover study to assess VCN effects on measures of gait, balance, and cognition (**Figure 1**). Participants completing the initial receptor occupancy study were eligible to enroll in this experiment. Participants were randomized 1:1 to one of two treatment sequences: placebo followed by VCN 0.5 mg b.i.d., or VCN followed by placebo. A statistician prepared the randomization list using permuted blocks with random block sizes. The list with randomization number and treatment allocation was sent to the research pharmacy and a blinded list of randomization numbers was sent to the study coordinator. After patient consent was completed and eligibility confirmed, the coordinator assigned the next randomization number to the participant, and sent a prescription with participant ID and randomization number to the research pharmacist who dispensed the appropriate study medication. To mask drug, VCN pills or placebo were encapsulated in gelatin sheaths. Participants received an initial 0.25 mg dose or equivalent placebo following baseline evaluations and were monitored for 4 hours after initial study medication administration with total daily dose or equivalent placebo escalated over the next 2 days.

**Figure 1:**
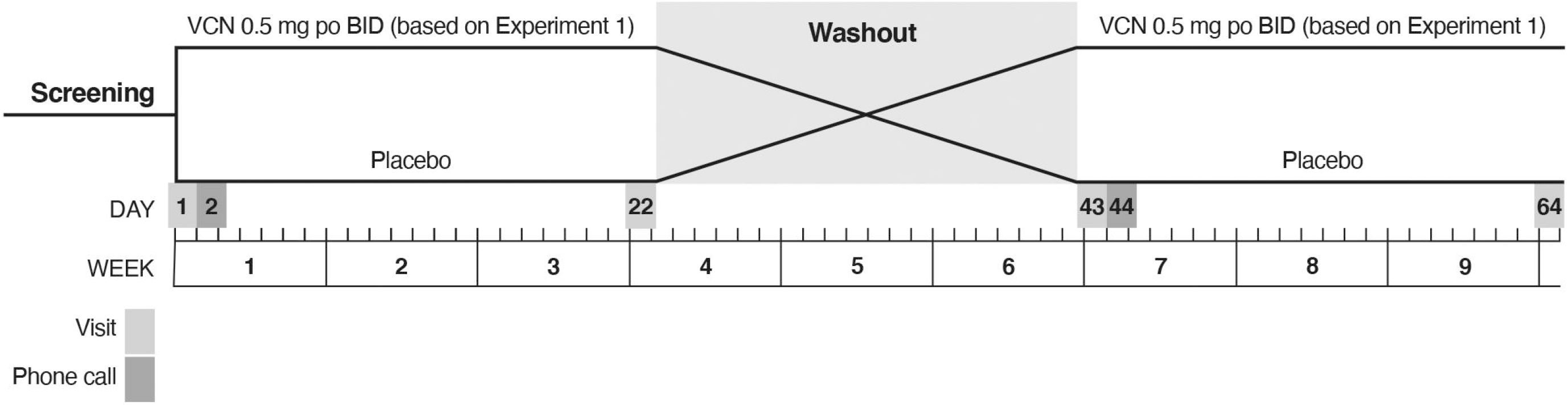
Design of Crossover Study.

Treatment periods were 3 weeks in duration and interrupted by 3 week washout periods. Participants underwent a standard evaluation at baseline, at the end of the first treatment period, at end of the washout period-beginning of the second treatment period, and at the end of the second treatment period (**Figure 1**). Outcome measures at the end of the VCN and placebo treatment periods were compared to assess VCN effects. The standard evaluation was a battery of motor, cognitive, and behavioral measures (see below). We *a priori* selected a measure of gait performance, normal pace gait speed, and a measure of postural stability, Mancini et al.’s JERK, as co-primary endpoints.^29,30^ JERK is the time based derivative of lower trunk accelerations during standing spontaneous sway. JERK was chosen because it tracks postural instability in PD.^30^ Normal pace gait speed was chosen as prior studies indicated that neocortical cholinergic denervation is associated with slower gait speed in PD.^8^ We hypothesized that VCN treated participants would ambulate faster and that VCN treatment would reduce JERK. Gait analysis was performed on an 8 meter GAITRite pressure sensitive walkway (CIR Systems, Inc.) and standard parameters were analyzed using ProtoKinetics Movement Analysis Software (GAITRite version 5.09C; ProtoKinetics, LLC). Gait assessments were repeated with a dual-task protocol in which participants counted backwards by three starting at a random number (under 100) provided by the examiner. Postural stability was assessed with the Ambulatory Parkinson’s Disease Monitoring (APDM) wearable sensor system (APDM Wearable Technologies, Inc.) using the iSWAY protocol, with participants standing on a foam pad with eyes open and eyes closed. Standard postural measures, including JERK, were assessed and calculated using the manufacturer’s software (Mobility Lab Version 1). We used the APDM system’s iTUG (Timed Up and Go) protocol to collect additional exploratory data. For cognition, we used a general cognitive measure, the Montreal Cognitive Assessment (MoCA) and selected tests to examine major cognitive domains but focusing on attention and executive function.

### Motor Assessments

Movement Disorder Society Unified Parkinson’s Disease Rating Scale, part III (MD-UPDRSIII; “on” state); MDS-UPDRSIII postural instability and gait subscore (PIGD) subscale score (sum of items 3.1, 3.9-3.13); Gait Speed (normal pace); Gait Speed (fast pace); Gait Speed (normal pace - dual task); Gait Speed (fast pace – dual task); Postural stability measures – mean sway velocity, JERK, root mean square sway distance (RMS), and dual support time during gait. To assess the effects of attentional loading, normal pace and fast pace gait were performed under dual task conditions. Differences in gait speed between dual task and no dual task conditions are a measure of the attentional burden imposed by the dual task. To assess the effects of VCN on this aspect of gait performance, we compared the differences between no dual task and dual task gait speed between VCN and placebo treatment periods. We also computed the Dual Task Cost (DTC; dual task gait speed minus no dual task gait speed divided by no dual task gait speed multiplied by 100), a standard metric of the attentional burden imposed by distractors during gait performance.^31^

### Cognitive Assessments

MoCA; Wechsler Adult Intelligence Scale-III Digit Symbol modalities test; California Verbal Learning Test (CVLT) short term memory test; CVLT long term memory test; CVLT recognition test; Delis-Kaplan Executive Function System (D-KEFS) Stroop III; D-KEFS sorting total; D-KEFS verbal fluency letters total; D-KEFS verbal fluency animals; D-KEFS Trail Making Test 4; Judgment of Line Orientation (JOLO) test. We assessed attentional function with a Sustained Attention Test (SAT), established to reflect CNS cholinergic systems function in humans.^32,33,34^ The SAT is performed with 2 conditions; without (SAT) and with a distractor (dSAT). SAT and dSAT results are reported as the vigilance index, a measure that corrects estimates of accurate detection with penalties for false detections and not confounded by errors of omission.^35^

### Behavioral Assessments

Geriatric Depression Score (GDS) and Columbia-Suicide Severity Rating Scale (C-SSRS).

### Treatment Compliance Monitoring

To assess compliance, we measured plasma VCN levels at the ends of treatment periods. VCN concentrations were assessed by the University of Michigan College of Pharmacy Pharmacokinetics Core. Plasma samples were deproteinated with acetonitrile, extracts centrifuged at 3500 RPM for 10 minutes, and supernatants used for liquid chromatography-mass spectrometry/mass spectrometry (LC-MS/MS). Calibration curve with VCN concentrations from 2.5 ng/ml 250 ng/ml was highly linear (r=0.999). Assay accuracy and precision were evaluated at 5 ng/ml, 10 ng/ml, and 200 ng/ml (N=3). Accuracy was 106% or less and precision was 10% relative standard deviation or less.

### Statistical Plan

The sample size (planned initially at four participants per dosing group) for the VCN-α4β2^*^ nAChR occupancy study was based on logistical considerations. For the crossover study, we calculated that 33 participants would provide at least 80% power to detect within-patient treatment differences of 0.122 m/s in gait speed and −0.131 m^2^/s^5^ for JERK, assuming within-participant correlation of ≥ 0.64 and ≥ 0.72, respectively, using a paired t-test and a two-sided Type I error of 0.025 (Bonferroni adjustment for co-primary endpoints). This approach is conservative given our analysis method uses mixed effects models. Estimates for treatment differences were based on Bohnen *et al*. for normal pace gait speed and Mancini *et al*. for JERK.^8,29^

We conducted exploratory analyses to examine the distributions of outcomes under each treatment, as well as individual and mean profiles over time. Graphical approaches such as boxplots and scatterplots with linear or non-linear (e.g., loess) methods were used, allowing identification of outliers, linearity, and correlation of measurements within participant and across time. Log transformations were applied when outcome data did not appear normally distributed.

Descriptive statistics for efficacy and safety outcomes were provided for each dosing cohort in the VCN-α4β2^*^ nAChR occupancy study. For the crossover study, linear mixed models containing treatment sequence, treatment period, treatment group, and dependent-variable baseline value, with participant within treatment sequence as a random effect, were used for analysis of continuous outcomes. To compare differences between VCN and placebo, a test for carryover based on the sequence effect was conducted using patient with sequence as the error term. Results are presented as least squares (LS) mean and standard error (SE). The co-primary endpoints were tested at the 2-sided, 0.025, significance level. All other tests were based on a 2-sided significance level of 0.05; no adjustments for additional multiple comparisons. Hence, p-values should be interpreted in the context of hypothesis generation in this target engagement study. 95% CIs are reported to estimate the magnitude of treatment effects.

Primary and secondary efficacy endpoints were analyzed in all randomized participants (intention-to-treat [ITT] population). Secondary continuous endpoints were analyzed similarly to the primary endpoints. Categorical analyses were based on Gart’s test.^36^

Safety endpoints were analyzed in all randomized participants who received at least one dose of study medication. We included adverse events that occurred in the washout period with the treatment given in period one.

## Results

### Participants

Characteristics of the fifteen PD participants enrolled for the initial VCN-α4β2^*^ nAChR occupancy study are described in **Table 1a**. Ten participants completed this phase of the study; two participants received 0.25 mg VCN per day, three participants 0.25 mg bid VCN per day, three participants 0.5 mg bid per day, and two participants 1.0 mg bid per day. Of the five PD participants not completing the imaging substudy, one PD participant was unable to tolerate PET imaging, tracer synthesis failed in two PD participants, and two PD participants discontinued VCN before PET imaging could be attempted. To confirm that α4β2^*^ nAChR – VCN interactions were not grossly different in PD compared to normal brain, an additional ten control participants were studied with ascending doses of VCN and [^18^F]Flubatine PET in a protocol identical to that used for PD. All ten control participants completed the imaging study protocol. Data from one control participant were excluded because of suspected covert tobacco abuse. Four participants received 0.25 mg per day, three participants 0.25 mg po b.i.d. per day, and three participants 0.5 mg b.i.d. per day. Characteristics of participants for the VCN-α4β2^*^ nAChR occupancy studies are described in **Table 1a**.

**Table 1A:** VCN-α4β2^*^ nAChR Occupancy Study Participant Baseline Characteristics.

|  | **PD**<br>**Participant**<br>**N = 15** | **Control Participants**<br>**N=10** |
| --- | --- | --- |
| **Age (Years)** |  |  |
| Mean (SD) | 67.3 (5.20) | 66.4 (8.95) |
| Min, Max | 52, 73 | 50, 76 |
| **Male, n (%)** | 14 (93%) | 8 (80%) |
| **White, n (%)** | 14 (93%) | 10 (100%) |
| **Age at Diagnosis (Years)** |  |  |
| Mean (SD) | 61.4 (5.69) | Not applicable |
| Min, Max | 51, 70 |  |
| **MDS-UPDRS III** |  |  |
| Mean (SD) | 31.9 (12.83) | 3.5 (1.84) |
| Min, Max | 9, 57 | 1, 7 |
| **GDS** |  |  |
| Mean (SD) | 3.2 (3.75) | 1.6 (2.01) |
| Min, Max | 0, 12 | 0, 6 |
| **MoCA** |  |  |
| Mean (SD) | 25.5 (1.73) | 26.8 (2.30) |
| Min, Max | 23, 29 | 23, 30 |

Characteristics of the crossover study participants are shown in **Table 1b**.

**Table 1B:**
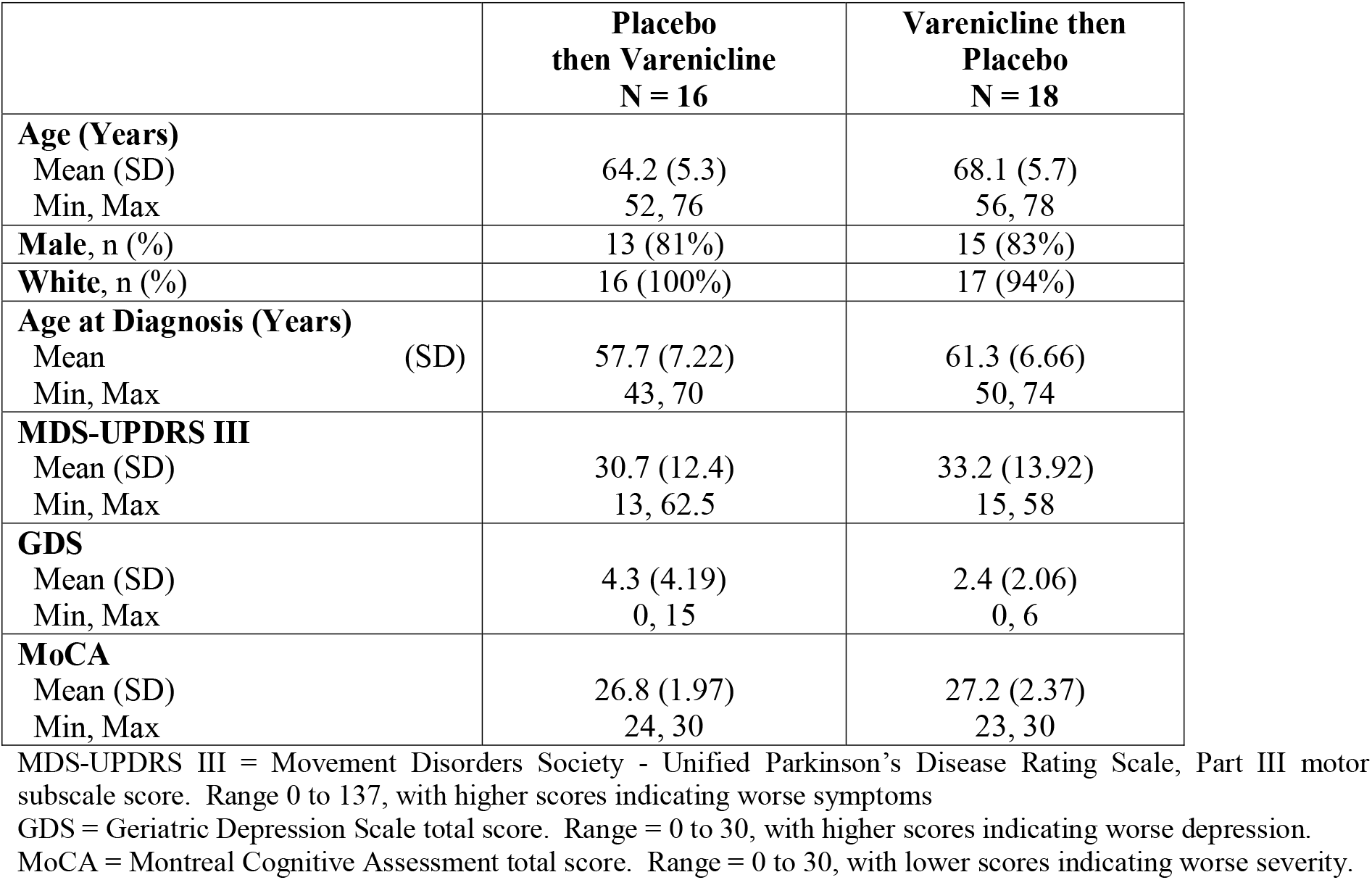
Crossover Study Participant Baseline Characteristics.

### VCN-α4β2^*^ nAChR Occupancy Study

VCN displacement of thalamic [^18^F]Flubatine binding to α4β2^*^ nAChRs is reported, the region with the highest [^18^F]Flubatine binding (**Figure 2**). Analysis of other regions gave very similar results (data not shown). The lowest daily dose of VCN, 0.25 mg per day, produced significant receptor occupancy. There was little evidence of a dose-response relationship with all VCN doses producing 60-70% occupancy of α4β2^*^ nAChRs. Results in control participants were very similar (data not shown). As 0.5 mg p.o. b.i.d. produced approximately the same α4β2^*^ nAChR occupancy as 1.0 mg p.o. b.i.d., 0.5 mg p.o. b.i.d. was chosen as the dose for the crossover study.

**Figure 2:**
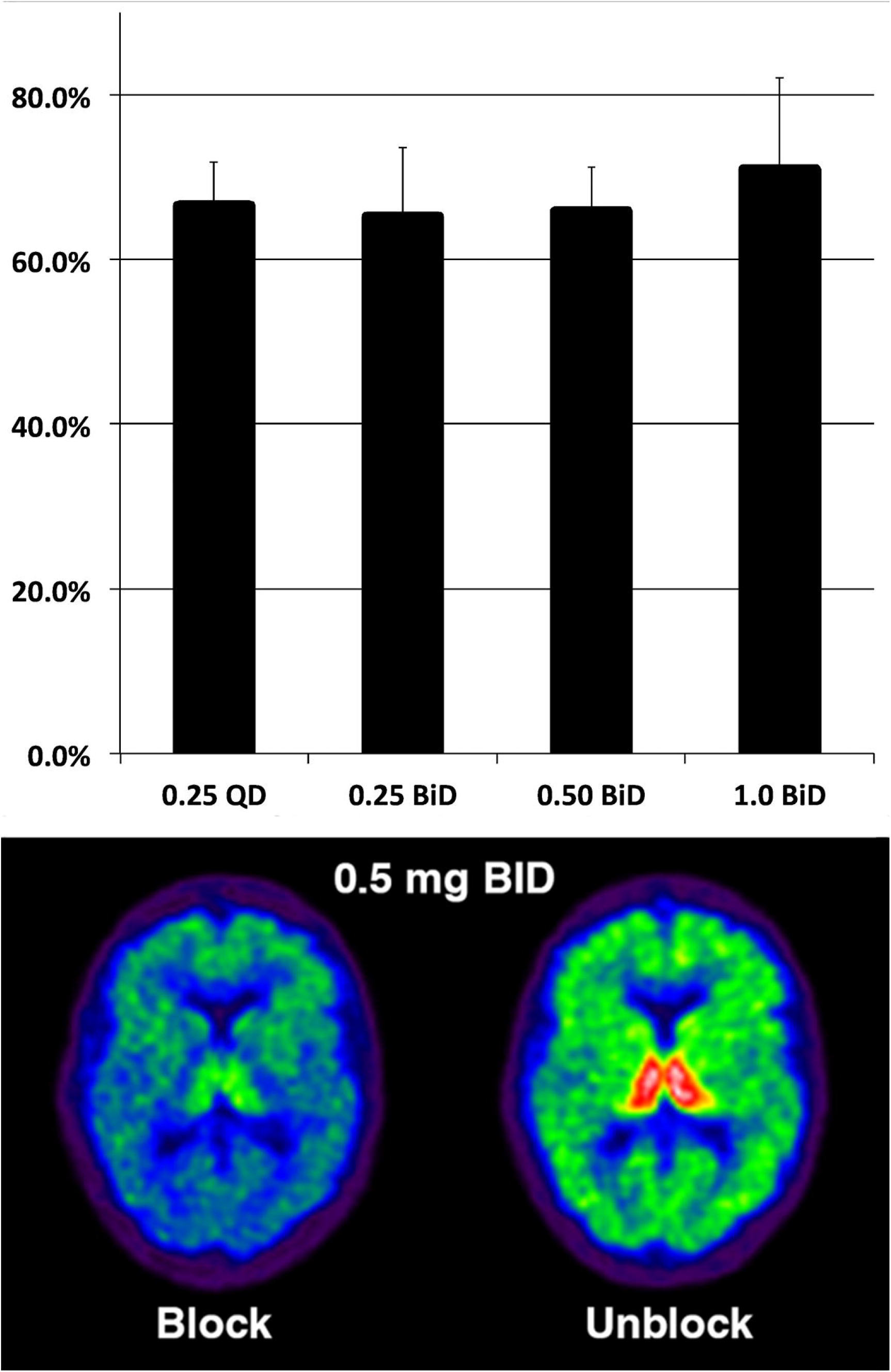
Varenicline Occupancy of α4β2^*^ nAChRs. Top panel is dose-response relationship between daily oral dose and estimated per cent receptor occupancy (mean and standard deviation). X-axis units are mg. Bottom panel is parametric images of a single participant on and off 0.5 mg po b.i.d.

### Crossover Study

#### Intention-to-Treat and Treatment Compliant Participants Analyses

The primary analyses of the crossover study used the ITT population. Secondary analyses were conducted using identical statistical methods in participants who were compliant with study treatment. In this secondary analysis, we excluded participants without evidence of significant increases in VCN plasma levels between placebo and VCN treatment periods to define the treatment compliant participants. Safety analyses are based on all participants.

#### Safety

We enrolled 34 PD participants. There was 1 drop-out for reasons unrelated to the study (withdrawn 4 days into period 2 while on VCN). Among the 34 participants, there were 56 adverse events (serious and non-serious) in 22 (65%) participants (**Table 2**). There were more adverse events in the VCN periods than placebo periods: 17 participants on VCN experienced 35 AEs, while 8 participants on placebo experienced 18 AEs. These were largely expected adverse events such as nausea and insomnia. There were 2 serious adverse events; one in each treatment period and neither related to VCN treatment. Two participants required dose reductions to 0.5 mg p.o. per day and 1 participant to 0.25 p.o. mg per day – 0.25 mg p.o. b.i.d. for study completion. As the VCN-α4β2^*^ nAChR occupancy experiment demonstrated substantial nAChR occupancy at the lowest VCN dose, these participants are included in all analyses.

**Table 2:** Crossover Study Adverse Events (AEs)

|  | **Placebo**<br>**N=34** | **Varenicline**<br>**N=34** |
| --- | --- | --- |
| **# Serious AEs** | 1 | 1 |
| **# Participants with $\geq 1$ Serious AE** | 1 | 1 |
| **# Non-Serious AEs<sup>a</sup>** | 18 | 35 |
| **# Participants with $\geq 1$ Non-Serious AE** | 8 | 17 |
| **Non-Serious AE Severity** |  |  |
| Mild | 15 | 29 |
| Moderate | 3 | 6 |
| **Non-Serious AE Relatedness** |  |  |
| Related | 4 | 16 |
| Unrelated | 14 | 19 |

<sup>a</sup>8 non-serious AEs occurred during washout, 6 in the placebo then varenicline arm and 2 in the varenicline then placebo arm; AEs during washout are categorized to the treatment in period 1 in this table; one subject randomized to varenicline then placebo had an unknown date of onset and thus is not included in this table (but their non-serious AE is included in summaries in the text).

#### Motor Function Measures – ITT Population

Of the primary outcome measures, there was no statistically significant difference in JERK performance between VCN and placebo treatment periods (**Table 3**). For normal pace gait speed, VCN treatment was associated with statistically significant gait slowing, a result opposite to the hypothesized effect (**Table 3**). Analysis of secondary/exploratory measures returned disparate results. MDS-UPDRSIII scores modestly worsened during the VCN treatment periods (**Table 3**). The PIGD subscore was not significantly different between VCN and placebo periods. Other postural stability measures - mean sway velocity and RMS, were not significantly different between placebo and VCN treatment periods. There was no significant effect on no dual task dual support time. VCN did not significantly change normal pace dual task gait speed but significantly reduced the difference in normal pace gait speed between no dual task and dual task conditions. VCN treatment had no effect on gait speed under either fast pace conditions. For normal pace gait speed, VCN treatment produced a modest, but significant, reduction in DTC. There was no effect on the difference between fast pace gait speed under dual task and no dual task conditions or on fast pace gait speed DTC. VCN treatment had significant and predictable effects on normal pace cadence, stride length, and stride time. Consistent with lack of VCN effect on gait speed under the fast pace condition, there was no effect on fast pace cadence, stride length, stride time, or dual support time. iTUG measures showed no significant VCN effects (data not shown).

**Table 3:** Crossover Study Motor Outcomes. Intent-to-Treat Population (N=34)

|  | **Varenicline**<br>**LS Mean (SE)** | **Placebo**<br>**LS Mean (SE)** | **Treatment Difference**<br>**(95% CI)** | **P-value** |
| --- | --- | --- | --- | --- |
| **Primary Outcomes<sup>a</sup>** |  |  |  |  |
| **Gait Speed–Normal Pace–No Dual Task**<br>**(cm/s)** | 121.27 (1.36) | 124.89 (1.36) | **-3.62 (-5.92, -1.32)** | **0.003** |
| **JERK (m<sup>2</sup>/sec<sup>5</sup>)** | 0.97 (0.20) | 1.04 (0.20) | -0.07 (-0.44, 0.31) | 0.73 |
| **Secondary Outcomes** |  |  |  |  |
| **MDS-UPDRS III** |  |  |  |  |
| **Total** | 31.90 (1.15) | 28.77 (1.13) | **3.13 (0.45, 5.80)** | **0.02** |
| **PIGD Subscore** | 4.41 (0.32) | 4.89 (0.32) | -0.48 (-1.32, 0.36) | 0.25 |
| **Gait** |  |  |  |  |
| **Gait Speed–Normal Pace–Dual Task** | 114.99 (1.48) | 116.26 (1.48) | -1.27 (-4.03, 1.49) | 0.35 |
| **Gait Speed–Fast Pace–No Dual Task** | 148.92 (1.73) | 151.26 (1.73) | -2.34 (-5.71, 1.04) | 0.17 |
| **Gait Speed–Fast Pace–Dual Task** | 135.61 (2.08) | 137.76 (2.08) | -2.14 (-5.06, 0.78) | 0.14 |
| **Gait Speed–Normal Pace–Dual Task minus**<br>**Gait Speed–Normal Pace–No Dual Task** | -6.07 (1.11) | -9.10 (1.11) | **3.03 (0.53, 5.54)** | **0.02** |
| **Normal Pace Dual Task Cost** | -5.34 (0.91) | -7.59 (0.91) | **2.25 (0.28, 4.22)** | **0.03** |
| **Gait Speed–Fast Pace–Dual Task minus**<br>**Gait Speed–Fast Pace–No Dual Task** | -13.40 (1.37) | -13.81 (1.36) | 0.41 (-2.28, 3.11) | 0.76 |
| **Fast Pace Dual Task Cost** | -9.28 (0.95) | -9.26 (0.95) | 0.03 (-1.76, 1.71) | 0.98 |
| **Cadence–Normal Pace–No Dual Task** | 109.50 (0.58) | 111.73 (0.58) | **-2.23 (-3.63, -0.83)** | **0.003** |
| **Cadence–Fast Pace–No Dual Task** | 120.70 (0.85) | 121.97 (0.85) | -1.27 (-2.83, 0.28) | 0.10 |
| **Mean Stride Length–Normal Pace–No Dual**<br>**Task** | 132.58 (1.16) | 134.56 (1.16) | **-1.98 (-3.69, -0.26)** | **0.03** |
| **Mean Stride Length–Fast Pace–No Dual**<br>**Task** | 147.96 (1.18) | 148.75 (1.18) | -0.79 (-2.99, 1.41) | 0.47 |
| **%CV Stride Length–Normal Pace–No Dual Task** | 3.87 (0.23) | 3.67 (0.23) | 0.20 (-0.27, 0.66) | 0.39 |
| **%CV Stride Length–Fast Pace–No Dual Task** | 3.55 (0.22) | 3.41 (0.22) | 0.14 (-0.41, 0.68) | 0.61 |
| **Mean Stride Time–Normal Pace–No Dual Task** | 1.10 (0.01) | 1.08 (0.01) | **0.02 (0.008, 0.04)** | **0.004** |
| **Mean Stride Time–Fast Pace–No Dual Task** | 1.00 (0.01) | 0.99 (0.01) | 0.01 (-0.003, 0.02) | 0.12 |
| **%CV Stride Time–Normal Pace–No Dual Task** | 2.55 (0.58) | 3.36 (0.58) | -0.81 (-2.42, 0.80) | 0.31 |
| **%CV Stride Time–Fast Pace–No Dual Task** | 2.54 (0.15) | 2.63 (0.15) | -0.09 (-0.50, 0.32) | 0.67 |
| **Dual Support-Normal Pace-No Dual Task** | 0.37 (0.01) | 0.37 (0.01) | 0.006 (-0.005, 0.018) | 0.25 |
| **Dual Support-Fast Pace-No Dual Task** | 0.31 (0.01) | 0.31 (0.01) | 0.002 (-0.009, 0.013) | 0.70 |
| **Mean Sway Velocity** | 0.31 (0.03) | 0.26 (0.03) | 0.05 (-0.05, 0.15) | 0.30 |
| **Sway RMS** | 0.15 (0.01) | 0.14 (0.01) | 0.01 (-0.02, 0.04) | 0.43 |

LS = least squares; JERK = time-based derivative of lower trunk accelerations; PIGD = postural instability and gait disorder; RMS = root mean square.
Estimates (least squares or adjusted means and standard errors) and p-values obtained from linear mixed model containing treatment sequence, treatment period, treatment group, and dependent-variable baseline value, with participant within treatment sequence as a random effect. Postural measures (JERK; Mean Sway Velocity; RMS) analysis based on a model with baseline eyes open-foam pad covariate and baseline eyes closed-foam pad covariate.
<sup>a</sup>Nominal alpha = 0.025 owing to Bonferroni correction for two co-primary endpoints.

#### Cognitive Measures – ITT Population

The SAT, a measure of attentional function that reflects CNS cholinergic functions, showed positive effects of VCN treatment (**Table 4**). There was no significant effect of VCN on dSAT performance, which was poor in both VCN treatment and placebo periods. Neither the MoCA, nor any conventional cognitive domain specific measures showed significant effects of VCN treatment (**Table 4**).

**Table 4:** Crossover Study Cognitive and Behavioral Outcomes. Intent-to-Treat Population (N=34)

|  | **Varenicline**<br>**LS Mean (SE)** | **Placebo**<br>**LS Mean (SE)** | **Treatment Difference**<br>**(95% CI)** | **P-value** |
| --- | --- | --- | --- | --- |
| **Cognitive Measures Reflecting**<br>**Cholinergic Signaling** |  |  |  |  |
| SAT<sup>a</sup> | 0.73 (0.06) | 0.66 (0.06) | 0.076 (0.0073, 0.14) | 0.03 |
| dSAT<sup>a</sup> | 0.49 (0.06) | 0.43 (0.06) | 0.058 (-0.03, 0.15) | 0.18 |
| **Conventional Cognitive Measures** |  |  |  |  |
| MoCA | 26.83 (0.29) | 27.23 (0.29) | -0.40 (-1.22, 0.43) | 0.34 |
| WAIS-III Digit Symbol | 62.38 (1.29) | 62.34 (1.28) | 0.04(-1.92, 2.00) | 0.97 |
| CVLT STM | 12.03 (0.32) | 12.36 (0.32) | -0.33 (-1.14, 0.48) | 0.41 |
| CVLT LTM | 12.44 (0.30) | 12.80 (0.30) | -0.36 (-0.77, 0.04) | 0.07 |
| CVLT recognition | 15.56 (0.13) | 15.47 (0.13) | 0.09 (-0.11, 0.30) | 0.36 |
| D-KEFS Stroop III (interference) | 112.52 (3.42) | 115.31 (3.37) | -2.79 (-12.06, 6.47) | 0.54 |
| JOLO | 25.84 (0.48) | 24.48 (0.48) | 0.36 (-0.79, 1.52) | 0.53 |
| D-KEFS Sorting Total | 84.55 (1.57) | 85.63 (1.55) | -1.08 (-4.66, 2.49) | 0.54 |
| D-KEFS Verbal Fluency - Letters Total | 43.53 (1.05) | 41.69 (1.04) | 1.83 (-0.61, 4.27) | 0.14 |
| D-KEFS Verbal Fluency - Animal | 18.19 (0.72) | 18.07 (0.71) | 0.12 (-1.33, 1.56) | 0.87 |
| Trail Making Test TMT4 | 103.19 (8.12) | 103.38 (8.03) | -0.19 (-16.37, 15.98) | 0.98 |
| **Behavioral** |  |  |  |  |
| GDS | 3.56 (0.28) | 3.31 (0.27) | 0.25 (-0.33, 0.84) | 0.38 |

LS = least squares; MoCA = Montreal cognitive assessment; WAIS = Wechsler adult intelligence scale; CVLT = California verbal learning test; Delis-Kaplan executive function system; SAT = Sustained attention test; dSAT = Distractor sustained attention test; GDS = Geriatric depression scale.
Estimates (least squares or adjusted means and standard errors) and p-values obtained from linear mixed model containing treatment sequence, treatment period, treatment group, and dependent-variable baseline value, with participant within treatment sequence as a random effect.
<sup>a</sup> Estimates (least squares or adjusted means and standard errors) and p-values obtained from linear mixed model containing treatment sequence, treatment period, and treatment group, with participant within treatment sequence as a random effect.

#### Behavioral Measures – ITT Population

VCN treatment had no effects on GDS (**Table 4**) or C-SSRS scores (data not shown).

#### Analysis of Treatment Compliant Population

We excluded the participant who dropped out prior to study completion. Four participants exhibited VCN levels below the level of quantification at the end of both treatment periods, suggesting non-compliance. All analyses of motor, cognitive, and behavioral measures were repeated for the 28 compliant participants (**Tables 5** and **6**). Results were essentially identical to those of the ITT population. For the primary endpoints, VCN was associated with significantly slower normal pace gait speed and no significant effect on JERK. VCN significantly improved SAT performance. There was a similar reduction in the difference between dual task normal pace gait performance and no dual task normal pace gait performance in the compliant population (p=0.06) and on normal pace DTC (p=0.07), however, these were not statistically significant in this smaller sample (**Table 5**). The magnitude of the mean VCN effect on the difference between dual task normal pace gait speed and no dual task normal pace gait speed, and normal pace DTC in the compliant population were very similar to that seen in the ITT population (**Tables 3** & **5**).

**Table 5:** Crossover Study Motor Outcomes. Compliant Participants (n=28)

|  | **Varenicline**<br>**LS Mean (SE)** | **Placebo**<br>**LS Mean (SE)** | **Treatment Difference**<br>**(95% CI)** | **P-value** |
| --- | --- | --- | --- | --- |
| **Primary Outcomes<sup>a</sup>** |  |  |  |  |
| **Gait Speed–Normal Pace–No Dual Task (cm/s)** | 119.17 (1.54) | 122.06 (1.54) | **-2.90 (-5.48, -0.31)** | **0.03** |
| **JERK (m<sup>2</sup>/sec<sup>5</sup>)** | 1.03 (0.24) | 1.19 (0.24) | -0.16 (-0.61, 0.28) | 0.46 |
| **Secondary Outcomes** |  |  |  |  |
| **MDS-UPDRS III** |  |  |  |  |
| **Total** | 33.54 (1.29) | 30.72 (1.27) | 2.82 (-0.18, 5.81) | 0.06 |
| **PIGD Subscore** | 4.83 (0.36) | 5.40 (0.36) | -0.57 (-1.53, 0.39) | 0.23 |
| **Gait** |  |  |  |  |
| **Gait Speed-Normal Pace-Dual Task** | 112.62 (1.72) | 113.04 (1.71) | -0.42 (-3.19, 2.36) | 0.76 |
| **Gait Speed–Fast Pace–No Dual Task** | 145.67 (1.90) | 146.70 (1.89) | -1.03 (-4.92, 2.86) | 0.59 |
| **Gait Speed-Fast Pace-Dual Task** | 131.14 (2.29) | 132.56 (2.28) | -1.42 (-4.84, 2.01) | 0.40 |
| **Gait Speed–Normal Pace–Dual Task minus**<br>**Gait Speed–Normal Pace–No Dual Task** | -6.56 (1.28) | -9.24 (1.27) | 2.68 (-0.14, 5.49) | 0.06 |
| **Normal Pace Dual Task Cost** | -5.78 (1.04) | -7.85 (1.04) | 2.07 (-0.18, 4.32) | 0.07 |
| **Gait Speed–Fast Pace–Dual Task minus**<br>**Gait Speed–Fast Pace–No Dual Task** | -14.62 (1.44) | -14.48 (1.44) | -0.14 (-3.38, 3.11) | 0.93 |
| **Fast Pace Dual Task Cost** | -10.21 (1.03) | -9.90 (1.02) | -0.31 (-2.42, 1.80) | 0.76 |
| **Cadence–Normal Pace–No Dual Task** | 109.69 (0.66) | 111.87 (0.66) | **-2.17 (-3.87, -0.48)** | **0.01** |
| **Cadence–Fast Pace–No Dual Task** | 121.57 (0.91) | 121.93 (0.90) | -0.36 (-1.85, 1.13) | 0.62 |
| **Mean Stride Length–Normal Pace–No Dual Task** | 130.02 (1.34) | 131.51 (1.33) | -1.49 (-3.39, 0.41) | 0.12 |
| **Mean Stride Length–Fast Pace–No Dual Task** | 143.63 (1.33) | 144.40 (1.33) | -0.77 (-3.54, 2.01) | 0.57 |
| **%CV Stride Length–Normal Pace–No Dual Task** | 3.93 (0.26) | 3.81 (0.26) | 0.12 (-0.42, 0.66) | 0.64 |
| **%CV Stride Length–Fast Pace–No Dual Task** | 3.78 (0.26) | 3.54 (0.26) | 0.24 (-0.42, 0.89) | 0.47 |
| **Mean Stride Time–Normal Pace–No Dual Task** | 1.10 (0.01) | 1.07 (0.01) | **0.02 (0.005, 0.04)** | **0.02** |
| **Mean Stride Time–Fast Pace–No Dual Task** | 0.99 (0.01) | 0.99 (0.01) | 0.003 (-0.01, 0.02) | 0.67 |
| **%CV Stride Time–Normal Pace–No Dual Task** | 2.64 (0.68) | 3.61 (0.68) | -0.97 (-2.93, 0.98) | 0.31 |
| **%CV Stride Time–Fast Pace–No Dual Task** | 2.64 (0.16) | 2.64 (0.16) | -0.004 (-0.44, 0.44) | 0.99 |
| **Dual Support–Normal Pace–No Dual Task** | 0.37 (0.01) | 0.37 (0.01) | 0.003 (-0.009, 0.016) | 0.61 |
| **Dual Support–Fast Pace–No Dual Task** | 0.31 (0.01) | 0.32 (0.01) | -0.003 (-0.014, 0.008) | 0.52 |
| **Mean Sway Velocity** | 0.35 (0.04) | 0.27 (0.04) | 0.08 (-0.04, 0.19) | 0.16 |
| **Sway RMS** | 0.16 (0.01) | 0.14 (0.01) | 0.01 (-0.02, 0.04) | 0.48 |

LS = least squares; JERK = time-based derivative of lower trunk accelerations; PIGD = postural instability and gait disorder; RMS = root mean square.
Estimates (least squares or adjusted means and standard errors) and p-values obtained from linear mixed model containing treatment sequence, treatment period, treatment group, and dependent-variable baseline value, with participant within treatment sequence as a random effect.
<sup>a</sup>Nominal alpha = 0.025 owing to Bonferroni correction for two co-primary endpoints.

**Table 6:**
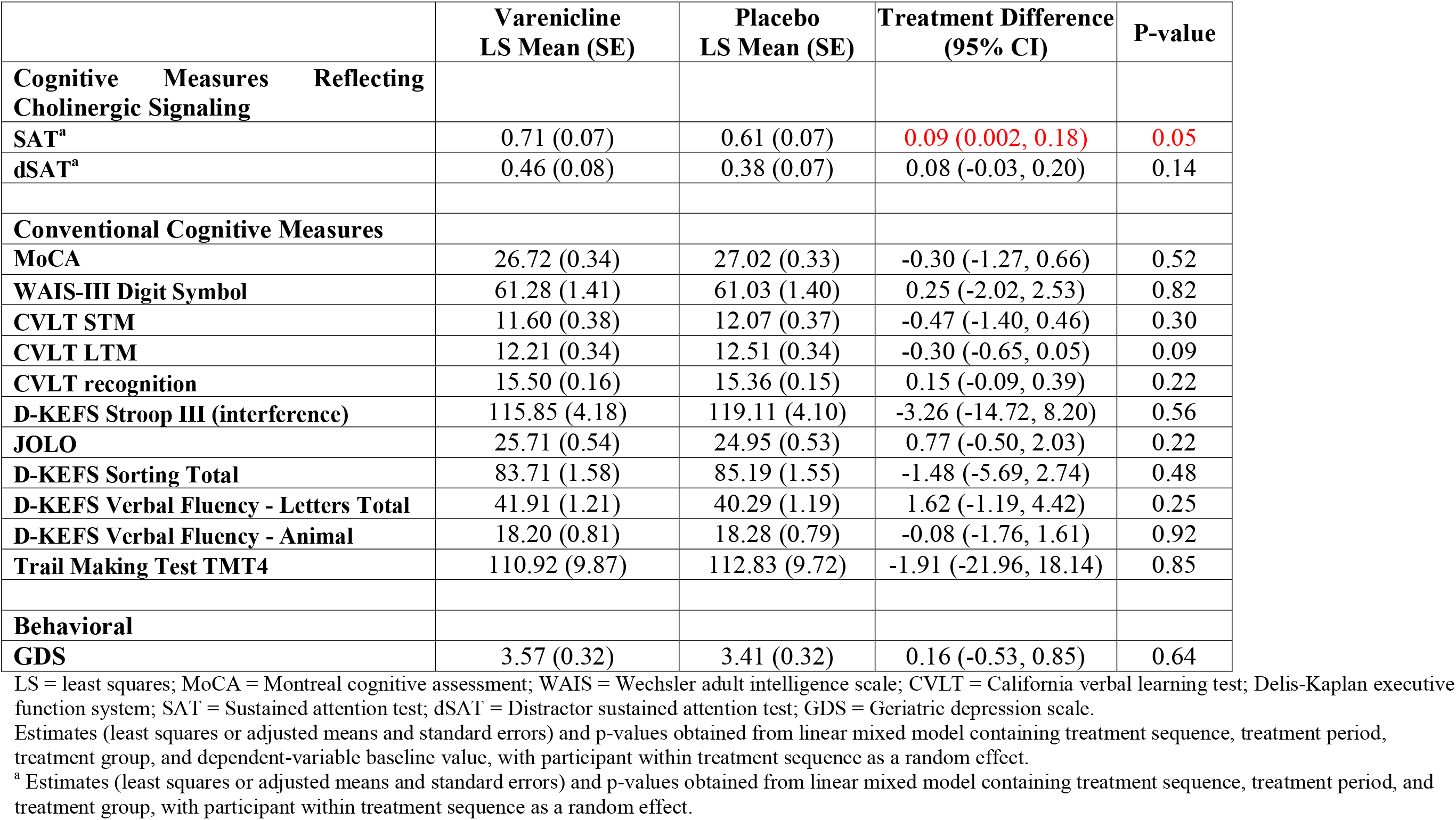
Crossover Study Cognitive and Behavioral Outcomes. Compliant Participants (n=28)

|  | **Varenicline**<br>**LS Mean (SE)** | **Placebo**<br>**LS Mean (SE)** | **Treatment Difference**<br>**(95% CI)** | **P-value** |
| --- | --- | --- | --- | --- |
| **Cognitive Measures Reflecting**<br>**Cholinergic Signaling** |  |  |  |  |
| **SAT<sup>a</sup>** | 0.71 (0.07) | 0.61 (0.07) | **0.09 (0.002, 0.18)** | **0.05** |
| **dSAT<sup>a</sup>** | 0.46 (0.08) | 0.38 (0.07) | 0.08 (-0.03, 0.20) | 0.14 |
| **Conventional Cognitive Measures** |  |  |  |  |
| **MoCA** | 26.72 (0.34) | 27.02 (0.33) | -0.30 (-1.27, 0.66) | 0.52 |
| **WAIS-III Digit Symbol** | 61.28 (1.41) | 61.03 (1.40) | 0.25 (-2.02, 2.53) | 0.82 |
| **CVLT STM** | 11.60 (0.38) | 12.07 (0.37) | -0.47 (-1.40, 0.46) | 0.30 |
| **CVLT LTM** | 12.21 (0.34) | 12.51 (0.34) | -0.30 (-0.65, 0.05) | 0.09 |
| **CVLT recognition** | 15.50 (0.16) | 15.36 (0.15) | 0.15 (-0.09, 0.39) | 0.22 |
| **D-KEFS Stroop III (interference)** | 115.85 (4.18) | 119.11 (4.10) | -3.26 (-14.72, 8.20) | 0.56 |
| **JOLO** | 25.71 (0.54) | 24.95 (0.53) | 0.77 (-0.50, 2.03) | 0.22 |
| **D-KEFS Sorting Total** | 83.71 (1.58) | 85.19 (1.55) | -1.48 (-5.69, 2.74) | 0.48 |
| **D-KEFS Verbal Fluency - Letters Total** | 41.91 (1.21) | 40.29 (1.19) | 1.62 (-1.19, 4.42) | 0.25 |
| **D-KEFS Verbal Fluency - Animal** | 18.20 (0.81) | 18.28 (0.79) | -0.08 (-1.76, 1.61) | 0.92 |
| **Trail Making Test TMT4** | 110.92 (9.87) | 112.83 (9.72) | -1.91 (-21.96, 18.14) | 0.85 |
| **Behavioral** |  |  |  |  |
| **GDS** | 3.57 (0.32) | 3.41 (0.32) | 0.16 (-0.53, 0.85) | 0.64 |

LS = least squares; MoCA = Montreal cognitive assessment; WAIS = Wechsler adult intelligence scale; CVLT = California verbal learning test; Delis-Kaplan executive function system; SAT = Sustained attention test; dSAT = Distractor sustained attention test; GDS = Geriatric depression scale.
Estimates (least squares or adjusted means and standard errors) and p-values obtained from linear mixed model containing treatment sequence, treatment period, treatment group, and dependent-variable baseline value, with participant within treatment sequence as a random effect.
<sup>a</sup> Estimates (least squares or adjusted means and standard errors) and p-values obtained from linear mixed model containing treatment sequence, treatment period, and treatment group, with participant within treatment sequence as a random effect.

#### VCN Levels

At the end of the placebo period, VCN levels were below the level of quantification for all participants, and mean VCN levels were 13.94 ng/ml (SD 7.91) at the end of the varenicline treatment periods. VCN concentrations at the end of the treatment period are consistent with previously reported pharmacokinetic data.^22^

## Discussion

We assessed potential VCN target engagement of α4β2^*^ nAChRs by assessing α4β2^*^ nAChR occupancy and behavioral effects. In our first experiment, we employed [^18^F]flubatine PET to establish that these VCN dose schedules produce significant α4β2^*^ nAChR occupancy without evidence of a dose-response relationship. These results are consistent with limited data suggesting that low oral VCN doses are effective for smoking cessation.^37^ We established that there is no gross difference in VCN-α4β2^*^ nAChR interactions in PD and control CNS. Our normal control results are similar to those obtained by Lotfipour *et al*., who used [^18^F]2-fluoro-3-[2(S)-2-azetidinylmethoxy]pyridine PET to demonstrate high α4β2^*^ nAChR occupancy in normal participants after a single, 0.5 mg, oral VCN dose.^38^ Our second experiment, using a daily dose based on receptor occupancy, was a placebo-controlled, double-masked crossover study. VCN had no effect on our primary measure of postural stability or other measures of postural stability studied. VCN was associated with slower normal pace gait speed and significantly narrowed the difference between normal pace gait speed under baseline and distracting dual task conditions with a significant effect on normal pace gait speed DTC. VCN had a significant positive effect on SAT performance, an attention measure directly linked to BFCC cholinergic functions, but no significant effects on conventional cognitive measures. The difference in SAT performance between VCN and placebo treatment periods is approximately the same as the difference in SAT performance between hypocholinergic and normocholinergic PD participants.^34^ Results were almost identical in analyses of ITT and treatment compliant participants. VCN narrowing of differences in dual task normal pace gait speed and no dual task normal pace gait speed, and normal pace DTC were not statistically significant in the compliant population, though of similar magnitude, likely reflecting the reduced sample size of the compliant population.

Our results complement and partly contradict those of Hall *et al*., who randomized 36 PD participants to VCN, 1 mg p.o. b.i.d., or placebo for an 8 week trial period.^38^ Participants were slightly older than our participants but comparable in PD severity and cognitive status. Their primary outcome measure was Berg Balance Scale (BBS) performance. Cognitive effects of VCN were assessed with the Mini-Mental State Examination (MMSE) and the Frontal Assessment Battery (FAB). There was no VCN effect with any of these measures. There are important methodological differences between this study and our work. Cholinergic systems are intact in many moderately advanced PD participants.^7,9^ It is likely that Hall *et al*. enrolled some participants with normal cholinergic systems, unlike our enrollment of participants with cortical cholinergic deficits. This difference may be important in the context of a partial agonist. In the presence of normal levels of endogenous agonists, partial agonists exhibit antagonist properties, potentially impairing cholinergic signaling. Likely most important is the difference in outcome measures. The BBS is a summary ordinal measure that does not quantify gait and balance measures and does not contain any distractors. The 6 item FAB contains 2 items (4 and 5) with attentional components, but there was no measure comparable to the SAT, a specific measure of attention reflecting cholinergic functions.

In terms of the selected primary endpoints, we found no effect on the measure of postural stability, JERK. We documented a significant effect on normal pace gait speed, though opposite to our hypothesis that nAChR stimulation would increase normal pace gait speed. It is plausible, however, that slower normal pace gait speed associated with VCN treatment might reflect greater attentiveness. In a rat model of variations in BFCC function, Kucinski et al. found that animals with better attentional capacity secondary to more robust BFCC function were more cautious during performance of an attentionally demanding gait task under single task conditions and less likely to fall under distracting conditions.^40^ The difference between normal pace gait speed at baseline and with a dual task distractor, showed a significant effect of VCN treatment and VCN treatment produced a significant improvement in DTC, a conventional measure of cognitive-motor integration. While there were no discernable effects of VCN on conventional measures of cognitive function, SAT performance, which reflects CNS cholinergic dysfunction in humans, including PD participants, showed a significant positive VCN effect. Similarly, Kucinski *et al*. showed that diminished vulnerability to falls is characteristic of high-attention capacity animals.^40^ Using the same model as the Kucinski *et al*., Paolone *et al*. demonstrated better SAT performance in animals with better BFCC function, and linked superior performance directly to α4β2^*^ nAChR stimulation.^41^ Our results are consistent also with those of Mocking *et al*., who showed that subacute oral VCN administration (0.5 mg/day for 3 days; then 1.0 mg/day for 4 days) in healthy participants improved working and declarative memory.^42^ Coupled with our [^18^F]flubatine PET data, we suggest that VCN treatment slowing of normal pace gait speed, narrowing of the difference between under baseline and distracting conditions, reduced DTC, the SAT results, constitute evidence of target engagement. In our analysis of exploratory/secondary endpoints, we did not correct for multiple comparisons, but it is notable that the only outcome measures with statistically significant results are plausibly related to attentional function.

We did not find any VCN effects under fast pace gait conditions. This may be because fast pace gait involves conscious focus on gait performance, strengthening attentional functions. Similarly, we did not find any VCN effect on dSAT performance, possibly due to floor effects. This conclusion is consistent with prior work indicating that the dSAT is very challenging for hypocholinergic PD participants.^34^ We did not find any effects on other postural control measures, mean sway velocity and RMS, studied, or on dual support time during gait. None of these postural control measures are directly linked to attentional functions or cholinergic deficits and may not be appropriate outcome measures to assess target engagement of attention. We noted a rise in MDS-UPDRSIII scores with VCN treatment. The modest magnitude of this effect is below the threshold of a minimally clinically important worsening in MDS-UPDRSIII scores.^43^

Our results highlight some of the difficulties involved with assessing interventions for DRT-refractory gait and balance disorders. Similar to the results of Hall *et al*., VCN did not have effects on conventional endpoints. One of our primary endpoint measures, no dual task normal pace gait speed, revealed a significant VCN effect. In our secondary analyses, we found positive effects of VCN treatment on normal pace gait performance when comparing dual task and no dual task conditions. Positive effects were found with the SAT, a measure that more closely reflects the cholinergic – attentional deficits that are likely major contributors to DRT-refractory gait and balance disorders. Targeting α4β2^*^ nAChRs may be a viable approach to mitigating this morbid PD feature. We suggest also that pursuing receptor subtype pharmacology, either for nAChRs or muscarinic cholinergic receptors, is more likely to be useful than non-specific approaches such as use of acetylcholinesterase inhibitors.^44^ Even in our hypocholinergic PD participants, there are regions with relatively preserved cholinergic innervation.^9^ As VCN is a partial agonist, it might impede normal cholinergic neurotransmission in these regions. Evaluation of full α4β2^*^ nAChR agonists may be worthwhile.

While we focus on the role of α4β2^*^ nAChRs in the BFCC system, α4β2^*^ nAChRs are widely distributed in the CNS (**Figure 2**), including high striatal expression where they are located on striatal afferent terminals and likely mediate some of the effects of striatal cholinergic interneurons.^45^ Activation of α4β2^*^ nAChRs on nigrostriatal dopaminergic terminals appears to enhance dopamine release. VCN effects could be mediated in part by α4β2^*^ nAChR stimulation in the striatum and other regions, though α4β2^*^ nAChR agonists do not have detectable motor effects in non-human primate models of PD.^45^

Future intervention studies for DRT-refractory gait-balance disorders will likely require laboratory-based measures that are both proxy measures of fall risk and permit efficient evaluation of target engagement. Future studies may also benefit from objective measures of fall risk derived from use of wearable sensors during daily life. Our experience suggests that outcome measures tied closely to underlying pathophysiologic mechanisms will be more robust biomarkers of target engagement.

## Data Availability

Data is available on request.

## Acknowledgments

The authors thank the study participants, and the staff of the Functional Neuroimaging, Cognition, and Mobility Laboratory and the PET Center. Supported by P50 NS091856, the Michael J. Fox Foundation, the Parkinson’s Foundation, and R21 NS114749.

## Author Contributions

Study Conception and Design: RLA, MLTMM, NIB, CS, MS, WTD

Acquisition and Analysis of Data: RLA, MLTMM, NIB, CS, AS, KK, CL, RAK, WTD

Drafting and Manuscript Review: RLA, MLTMM, NIB, CS, MS, AS, CL, KK, RAK, WTD

## Potential Conflicts of Interest

The authors have no conflicts of interest to declare.

## Notes

### Competing Interest Statement

The authors have declared no competing interest.

### Clinical Trial

NCT04403399; NCT02933372

### Funding Statement

Supported by: NIH-NINDS, Michael J. Fox Foundation, and the Parkinson's Foundation

### Author Declarations

University of Michigan IRBMED.

### Summary of Updates

1) Introduction and Discussion revised to emphasize target engagement focus of study. 2) Some additional analyses - addition of dual task cost.

